# Prevalence and Sociodemographic Covariates of Human Papillomavirus in the United States: Findings from the 2008 to 2020 National Inpatient Sample Database

**DOI:** 10.1101/2024.03.17.24304439

**Authors:** Emily Dantes, Mohammad Alfrad Nobel Bhuiyan, Margaret Bourg, Corey D. Smith, Md. Shenuarin Bhuiyan, Deborah G Smith

## Abstract

Human papillomavirus (HPV) infection is widespread worldwide, leading to a significant burden of HPV-related cancers. This study aimed to identify the prevalence of HPV infection in the United States based on a large nationwide inpatient sample database. We used the Nationwide Inpatient Sample (NIS) database from 2008 to 2020 and identified HPV cases using the International Classification of Diseases, Ninth (ICD-9) and Tenth Revision (ICD-10) codes. Pearson’s chi-square and independent-sample t-test were used for the analysis. The prevalence ratio was calculated using logistic regression models and the Cochran Armitage trend test to examine trends. A total of 47,506 HPV infections were identified from 2008 to 2020 among adults aged >18 years. Most patients were female (99.72%), with a higher prevalence among the 26-40 age group and white individuals. HPV infections were more prominent among low-income individuals and in the southern region of the United States. From 2008 to 2020, HPV increased from 6.76% to 10.91%. This study provides insights into the epidemiology of HPV infection in the United States. Understanding the trends and characteristics of HPV infection can contribute to public health interventions aimed at prevention and early detection.

## Introduction

Human papillomavirus (HPV) infection is widespread worldwide, leading to a significant burden of HPV-related cancers (1, 2). In the United States, HPV is the most common sexually transmitted infection, and it is estimated that 14 million Americans were newly infected with HPV in 2018 (3, 4, 5). Nearly all sexually active adults are infected at some point during their lives, although they may be unaware of their infection (5). While most HPV infections are asymptomatic and resolve spontaneously, long-lasting infections can progress into genital warts, as well as oropharyngeal and anogenital cancers (including anal, penile, vaginal, vulvar, and cervical) (2, 5).

More than 200 distinct types of HPV have been identified, with types falling into a low-risk or high-risk group (4). Low-risk types rarely lead to symptoms or clinical disease. High-risk or oncogenic HPV types, similarly asymptomatic at the onset of infection, may develop into cancer years later if undetected and untreated. According to the National Cancer Institute, high-risk HPV types cause 3% of all cancers in women and 2% in men in the United States (6). From 2015 to 2019, about 47,199 new HPV-associated cancers occurred annually in the United States (5, 7). Among women, cervical cancer is the most common HPV-associated cancer (5). It is currently the fourth most common cancer in women worldwide (2). Among men, oropharyngeal cancers (cancers of the back of the throat, including the base of the tongue and tonsils) are the most common (5, 8, 9). The incidence of oropharyngeal cancer in the United States increased by approximately 1.94% per year from 2000-2015 and has been projected to increase continuously over the next thirty years (10).

Many risk factors for HPV infection have been identified, including having multiple sexual partners, sexual intercourse at a younger age (<15), long-term use of oral contraceptives, smoking, low socioeconomic status, age, and genetic factors (5). Risk factors related to lifestyle behaviors can be addressed with public health intervention, which is necessary to prevent new infections and cancer progression (11). One important intervention is HPV vaccination, which has been demonstrated to prevent up to 90% of HPV-related cancers (6). The American Cancer Society recommends routine HPV vaccination for all persons aged 9 to 12 years old, with catch-up vaccination for those under 26 who are not adequately vaccinated (12). However, immunization rates in the US are much lower than for other vaccines, with only 59% of adolescents aged 13 to 17 years old being up to date with HPV vaccination as of 2020 (12). In addition, vaccination does not protect against all types of HPV, nor does it protect against already-established HPV infection (12). Another important intervention is early detection through screening; however, not all types of HPV-associated diseases and cancer have screening recommendations. For instance, while pap tests and HPV co-tests are available to screen for HPV-related cervical cancer, there is no method recommended by the American Cancer Society or the U.S. Preventative Services Task Force to screen for other HPV-related cancers like oropharyngeal, anal, penile, vulvar, or vaginal cancer (13). Due to these limitations, it is important to understand the current epidemiology of HPV infection in the United States. This study aims to understand the trends and identify the prevalence of HPV infection in the United States from 2008 to 2020 based on NIS data.

## Material & Methods

We used the Healthcare Cost and Utilization Project (HCUP)’s Nationwide Inpatient Sample (NIS) database from 2008 to 2020 (accessed December 19, 2022) and identified HPV-related hospital discharge cases using the International Classification of Diseases, ninth (ICD-9) and tenth Revision (ICD-10) codes to conduct a retrospective study. NIS’s database is the largest inpatient database, which covers information from seven million hospital stays yearly from 45 states in the United States. This database contains data from 20% of hospital discharges from all hospitals in HCUP (14).

To improve the sample’s representativeness, accuracy, and stability of weighted national estimates, the NIS was changed in 2012 to include 20% of all hospital discharges in HCUP. Before 2012, the NIS included all discharges from 20% of hospitals in the HCUP (15). The NIS data are organized so that each observation in the sample relates to a unique hospitalization. The NIS database provides information on more than 100 clinical characteristics, including patient demographics (such as age, sex, race, and the median income for ZIP code), hospital characteristics (such as ownership, size, teaching status, census region, and division), primary and up to 24 secondary diagnoses, 15 procedures as administrative codes, and codes for disease severity. Additionally, because it contains data on several million hospitalizations each year, it enables the analysis of healthcare utilization for issues related to rare diseases treated primarily in an inpatient setting.

Because the NIS database is deidentified, the Institutional Review Board (IRB) waived the IRB application. Authors did not have access to information that could identify individual participants during or after data collection. We included adult patients (age: >18 years and older) of male and female sex with a primary or secondary diagnosis of HPV based on the International Classification of Diseases, Ninth (ICD-9) and Tenth revision (ICD-10) diagnosis code (ICD-9 = 795.05, 795.09, 796.75, 796.79, ICD-10 = R87.810, R87.820, R85.81, R85.82) (Appendix A).

When analyzing NIS hospital discharge data, we adhered to AHRQ’s suggestions for producing national estimates from NIS data. Data was weighted with care to account for the change in database structure that occurred in 2012. The hospital discharge trend weight (TRENDWT) was used to weight data for 2012 and earlier, while regular discharge weight (DISCWT) was used to weight data for years after 2012. Descriptive statistics were used to characterize participant details. Continuous variables were summed for weighted and unweighted data, while mean and categorical variables were presented as frequencies, percentages, and 95% Confidence Intervals (CI). Design-based t-tests were used for continuous values, and design-based Chi-squared tests were applied for categorical values. The prevalence ratio was calculated using binomial logistic regression models. Linear regression models and the Cochran Armitage trend test were utilized to examine trends and contrast two groups. A p-value <0.05 was utilized to determine the statistical significance of the variables. All statistical tests were conducted using the free and open-source software R. (version 4.2.1).

## Results

We analyzed 395 million weighted hospital-discharged patients aged >18 or older from 2008-2020 NIS data. Overall, we found 47,506 hospital encounters with a primary or secondary diagnosis of HPV. Regarding the characteristics of the patients, most were females, 99.72%, 61.2% were between 26-40 years old, and 41.59% were Whites. HPV-related hospital discharges were more prominent among low-income individuals (35.33%) than high-income individuals (17.78%). The prevalence was higher in the southern United States (44.72%). In addition, among those patients with HPV, 19.3% had anemia, 15.4% were obese, and 10.2% were tobacco users (Table 1).

**Table 1.**
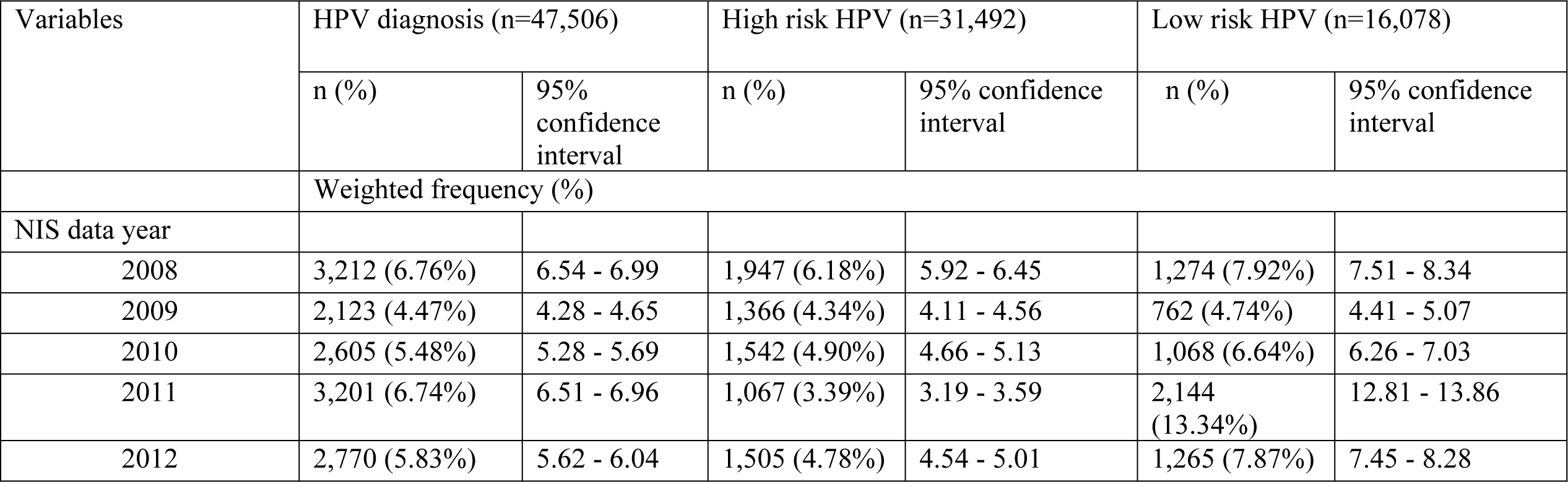
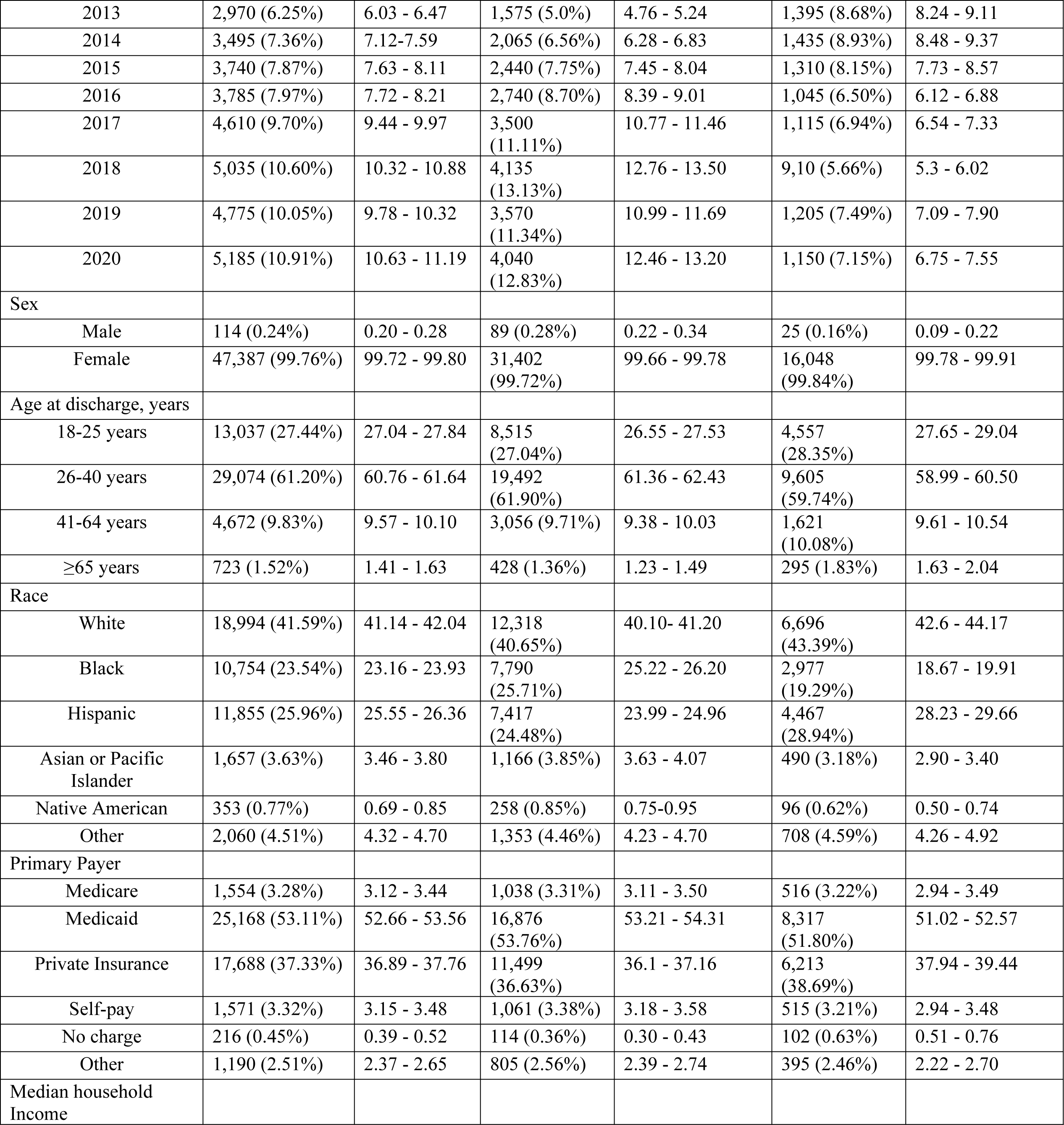
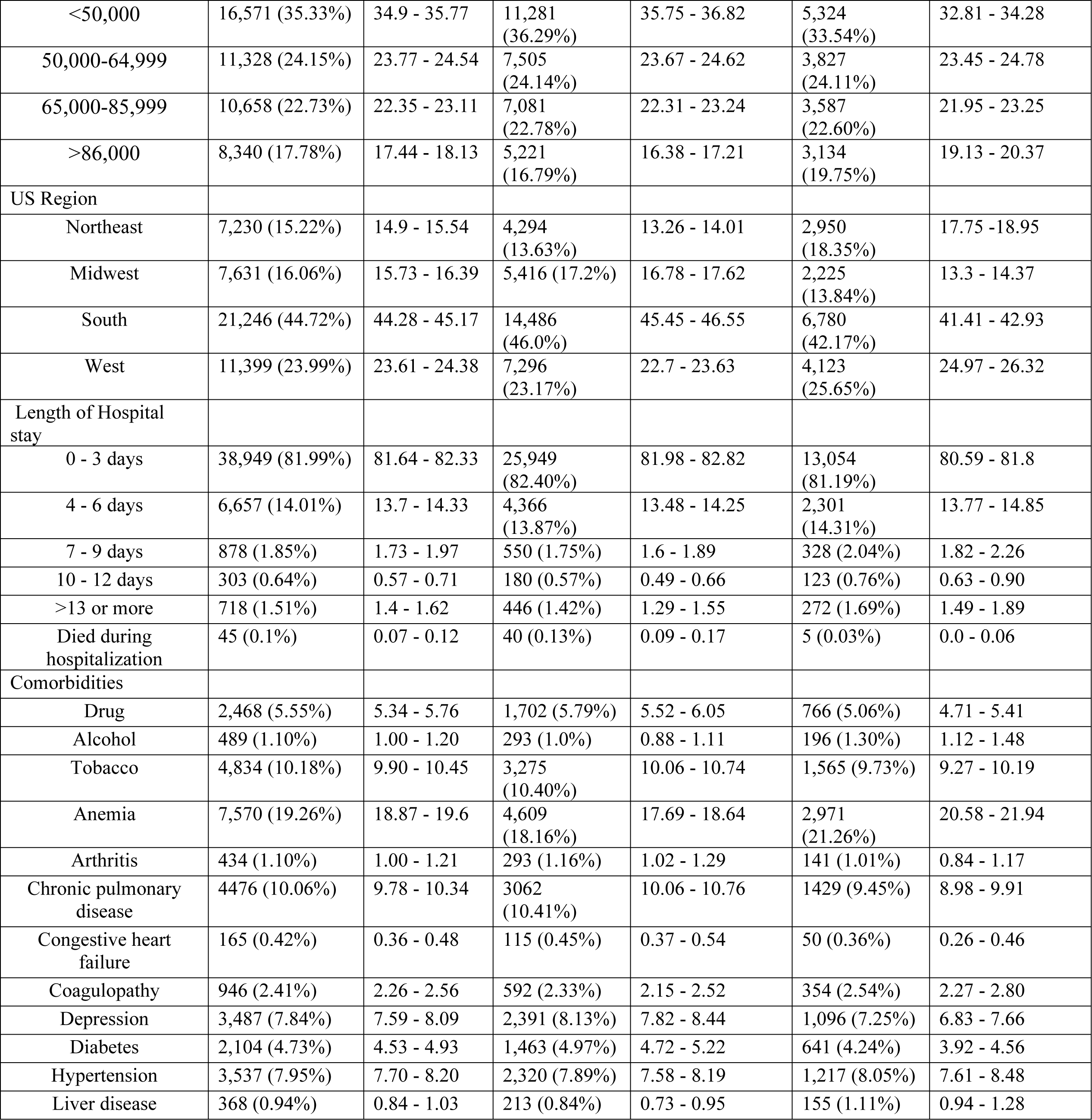
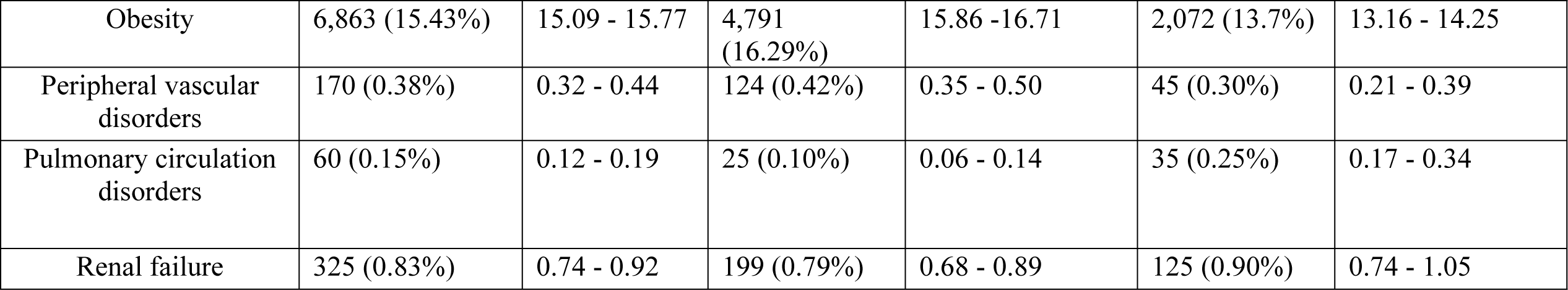
Demographic characteristics of patients with high-risk and low-risk HPV in the United States, 2008 - 2020.

Patients discharged from hospitals in the United States were 84 times more likely to have a primary or secondary diagnosis of HPV in 2020 compared to 2008 (95% CI: 1.67-2.03). The prevalence ratio of HPV was higher among females than males (PR: 295.27; 95% CI: 201.28 - 457.90). While the number of hospital discharges with an HPV-associated diagnosis was greatest among White individuals, the prevalence ratio of HPV was 398 times higher among Hispanics than White individuals (95% CI: 3.78-4.18), followed by Blacks (PR: 228; 95% CI: 2.45-2.72). The prevalence ratio was also higher for the patients with Medicaid (PR: 45.14; 95% CI: 40.35 – 50.70). With the regions, the prevalence of HPV was 45 times higher in the South compared to the Northeast (95% CI: 1.36-1.53) and 57 times higher in the West compared to the Northeast (95% CI: 1.47-1.68) (Table 2).

**Table 2.** Prevalence ratio for HPV-related patient discharges and sociodemographic covariates in the United States, 2008 – 2020.

| Variables | Prevalence Ratio<br>(Unadjusted) | 95% confidence interval |  | P-value |
| --- | --- | --- | --- | --- |
|  |  | Lower | Upper |  |
| NIS data year |  |  |  | < 0.000 |
| 2008 | Reference |  |  |  |
| 2009 | 0.68 | 0.61 | 0.77 |  |
| 2010 | 0.8 | 0.72 | 0.90 |  |
| 2011 | 1 | 0.90 | 1.11 |  |
| 2012 | 0.89 | 0.79 | 0.99 |  |
| 2013 | 0.97 | 0.87 | 1.09 |  |
| 2014 | 1.15 | 1.04 | 1.28 |  |
| 2015 | 1.22 | 1.10 | 1.35 |  |
| 2016 | 1.23 | 1.11 | 1.37 |  |
| 2017 | 1.49 | 1.35 | 1.64 |  |
| 2018 | 1.64 | 1.49 | 1.80 |  |
| 2019 | 1.55 | 1.41 | 1.71 |  |
| 2020 | 1.84 | 1.67 | 2.03 |  |
| Sex |  |  |  | < 0.000 |
| Male | Reference |  |  |  |
| Female | 295.27 | 201.28 | 457.90 |  |
| Age (years) |  |  |  |  |
| 18 – 25 years | Reference |  |  |  |
| 26 – 40 years | 0.94 | 0.90 | 0.98 |  |
| 41-64 years | 0.08 | 0.08 | 0.09 |  |
| ≥65 years | 0.01 | 0.01 | 0.02 |  |
| Race |  |  |  | <0.000 |
| White | Reference |  |  |  |
| Black | 2.59 | 2.45 | 2.72 |  |
| Hispanic | 3.98 | 3.78 | 4.18 |  |
| Asian or Pacific Islander | 2.29 | 2.05 | 2.56 |  |
| Native American | 1.93 | 1.51 | 2.42 |  |
| Other | 2.45 | 2.21 | 2.70 |  |
| Primary Payer |  |  |  | <0.000 |
| Medicare | Reference |  |  |  |
| Medicaid | 45.14 | 40.35 | 50.70 |  |
| Private Insurance | 18.62 | 16.62 | 20.94 |  |
| Self-pay | 10.14 | 8.68 | 11.85 |  |
| No charge | 13.96 | 10.05 | 18.91 |  |
| Other | 11.46 | 9.69 | 13.55 |  |
| Median household Income, percentile |  |  |  | <0.000 |
| 0-25th | Reference |  |  |  |
| 26th-50th | 0.78 | 0.74 | 0.82 |  |
| 51th-75th | 0.81 | 0.76 | 0.85 |  |
| 76th-100th | 0.74 | 0.70 | 0.78 |  |
| US Region |  |  |  | <0.000 |
| Northeast | Reference |  |  |  |
| Midwest | 0.89 | 0.83 | 0.95 |  |
| South | 1.45 | 1.36 | 1.53 |  |
| West | 1.57 | 1.47 | 1.68 |  |
| Length of Hospital stay (days) |  |  |  | <0.000 |
| 0 - 3 days | Reference |  |  |  |
| 4 – 6 days | 0.40 | 0.38 | 0.42 |  |
| 7 – 9 days | 0.14 | 0.12 | 0.17 |  |
| 10 – 12 days | 0.11 | 0.09 | 0.14 |  |
| >13 days or more | 0.17 | 0.14 | 0.20 |  |
| Comorbidities |  |  |  | <0.000 |
| Drug | 1.20 | 1.10 | 1.32 |  |
| Alcohol | 0.22 | 0.18 | 0.26) |  |
| Tobacco | 0.42 | 0.39 | 0.44 |  |
| Anemia | 1.36 | 1.28 | 1.43 |  |
| Arthritis | 0.40 | 0.32 | 0.49 |  |
| Chronic pulmonary disease | 0.48 | 0.45 | 0.52 |  |
| Congestive heart failure | 0.04 | 0.03 | 0.06 |  |
| Coagulopathy | 0.54 | 0.47 | 0.62 |  |
| Depression | 0.67 | 0.62 | 0.73 |  |
| Diabetes | 0.16 | 0.14 | 0.17 |  |
| Hypertension | 0.09 | 0.08 | 0.09 |  |
| Liver disease | 0.30 | 0.24 | 0.38 |  |
| Obesity | 1.22 | 1.15 | 1.29 |  |
| Peripheral vascular disorders | 0.06 | 0.04 | 0.09 |  |
| Pulmonary circulation disorders | 0.08 | 0.04 | 0.13 |  |
| Renal failure | 0.06 | 0.05 | 0.08 |  |

### Hospital Discharges with a Primary or Secondary HPV Diagnosis

From 2008 to 2020, hospital discharges with a primary or secondary diagnosis of HPV increased from 6.76% to 10.91% (Figure 1). Our study also revealed notable differences in HPV trends across races, with white individuals having the highest number of HPV infections as compared to individuals from other races (p<0.001) (Figure 2). We observed a rising pattern among individuals aged between 26 and 40 years (p<0.001) (Figure 3). A greater number of HPV diagnoses was found in hospital encounters among females compared to males (p<0.001) (Figure 4).

**Figure 1.**
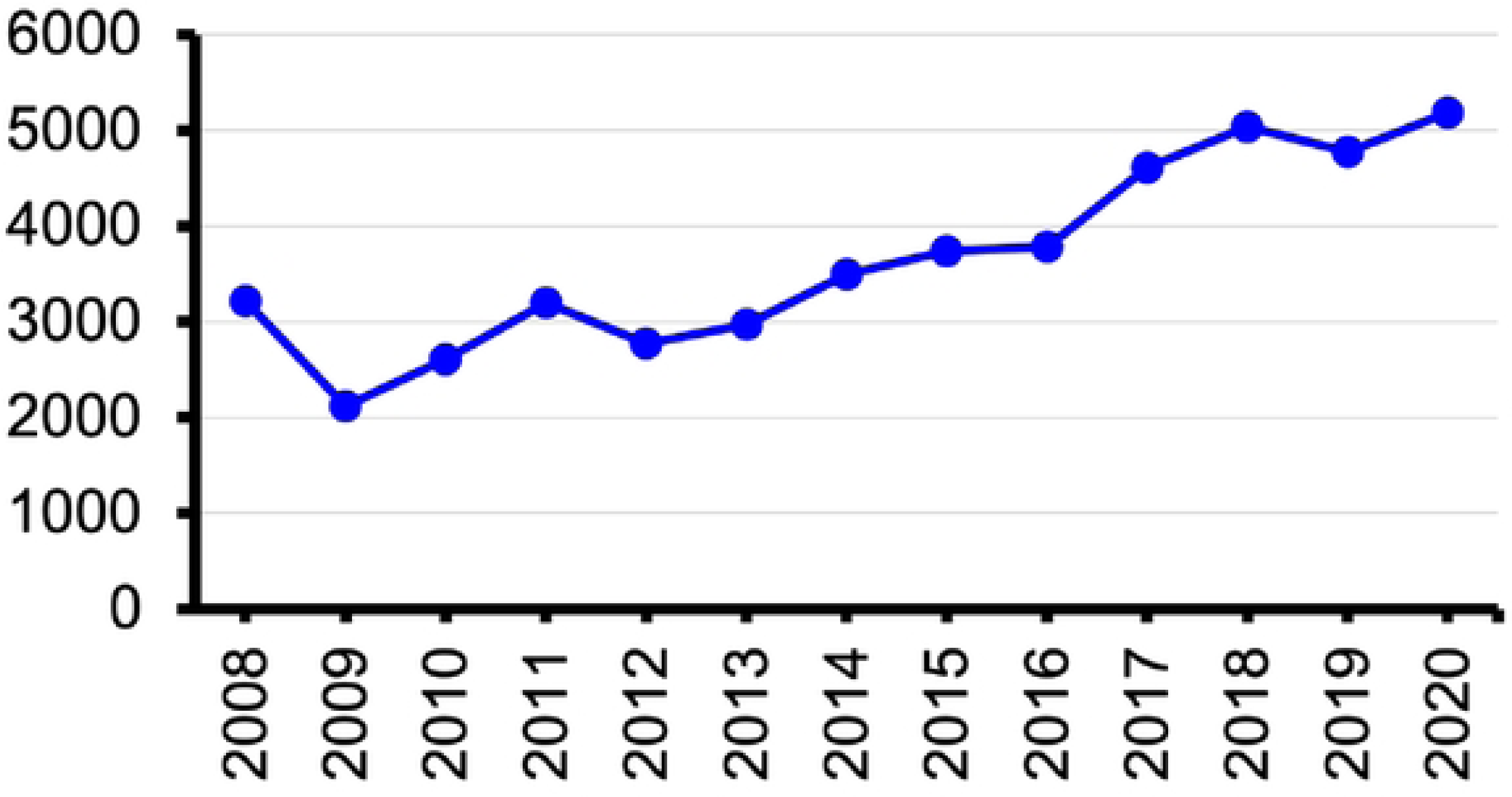
Trends in HPV-associated hospital discharge in the United States from 2008-2020

**Figure 2.**
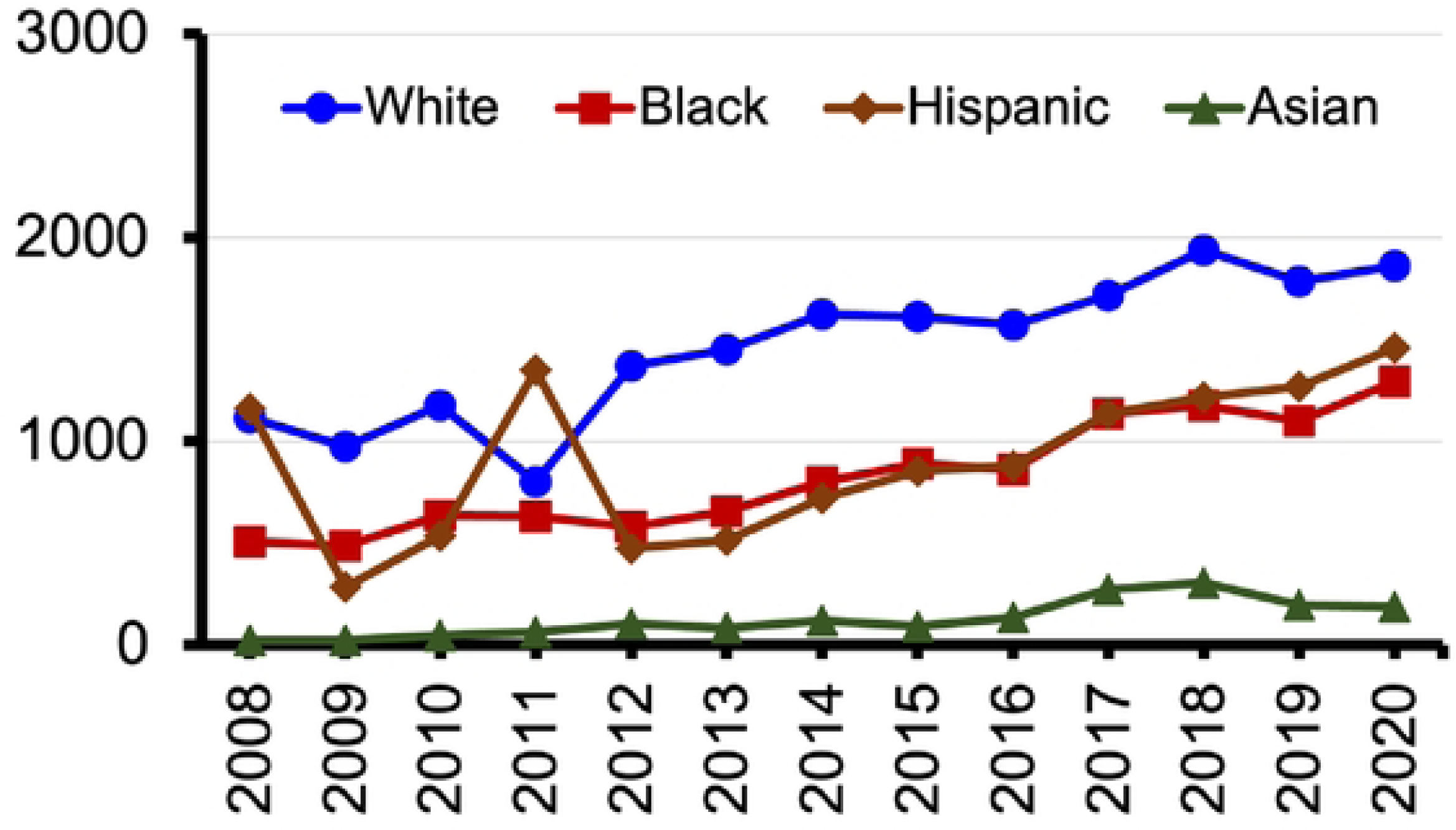
Trends in HPV-associated hospital discharge in the United States by race from 2008-2020

**Figure 3.**
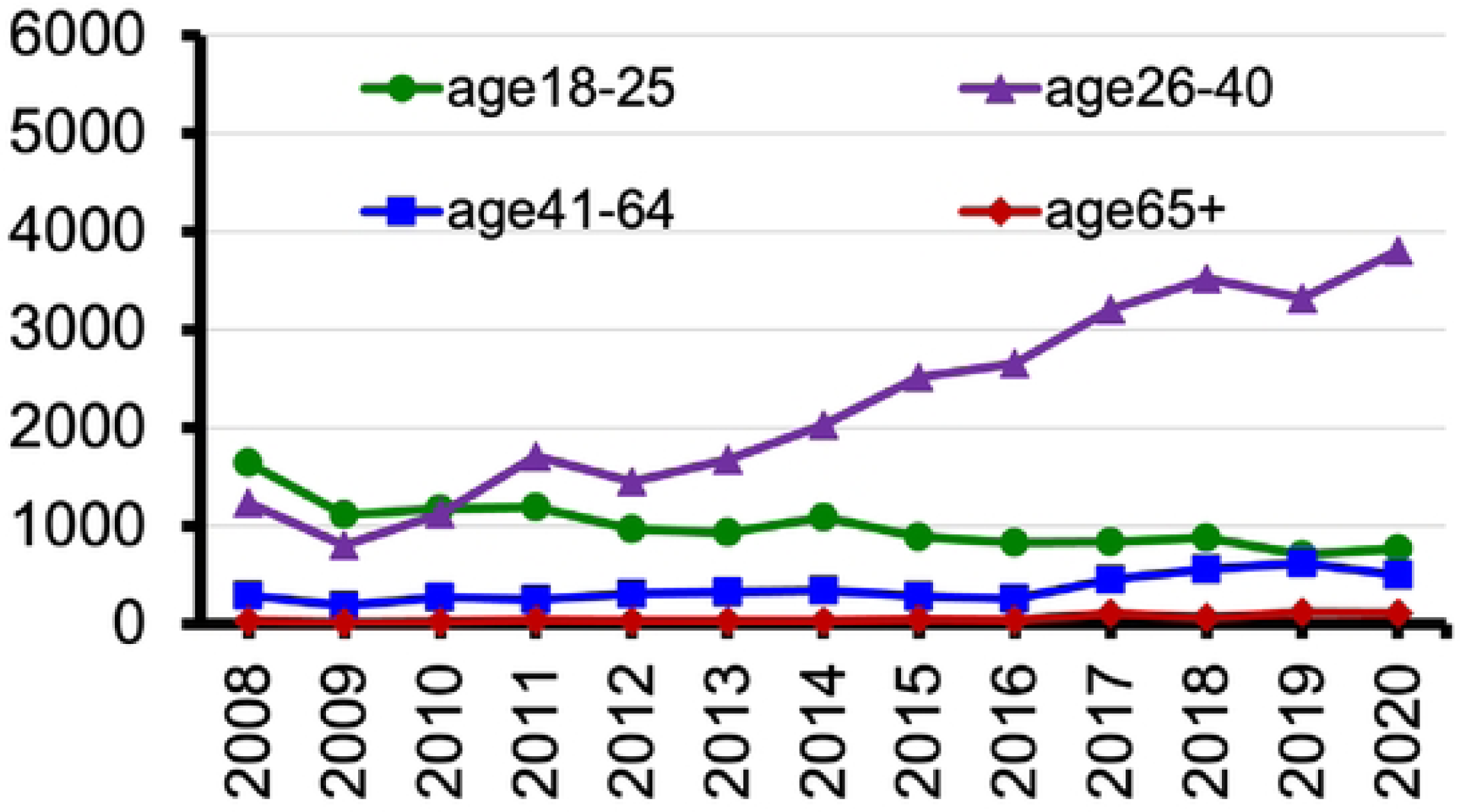
Trends in HPV-associated hospital discharge in the United States by age group from 2008-2020

**Figure 4.**
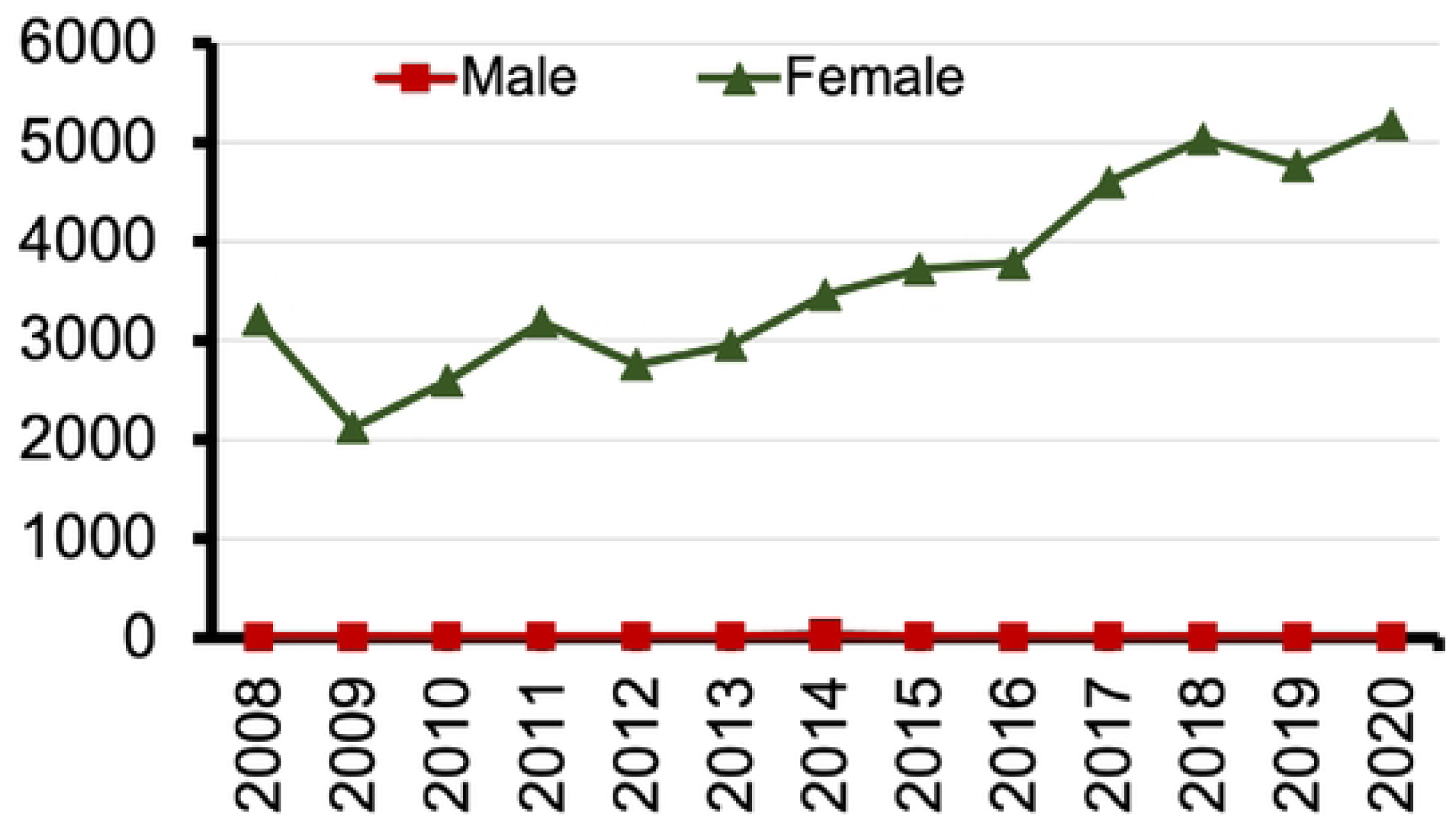
Trends in HPV-associated hospital discharge in the United States by sex from 2008-2020

## Discussion

Using hospital discharge registry data from NIS, we observed a significant increase in the incidence of a primary or secondary diagnosis of HPV in hospital discharges from 2008 to 2020 in the United States despite the introduction of HPV vaccination recommendations in 2006.

These increases were observed among women of younger age groups and the increase from 2008 to 2020 was highest in the 26 to 40 age group. These findings could indicate a greater ability to detect HPV and a higher emphasis being placed on HPV testing over time; however, the introduction of vaccination should theoretically have shown a decrease in the incidence of HPV-associated diagnoses over the course of 12 years despite increased detection.

These results follow the American Cancer Society’s findings that HPV vaccination rates remain low in the United States (12). This could be due to insufficient education of patients by healthcare providers about the risks of HPV infection or about the availability of HPV vaccination. This could also indicate a lack of public priority on annual wellness visits where these types of discussions may occur or distrust in the vaccine’s safety and efficacy. This study further supports the need to adopt evidence-based HPV vaccination programs, such as those listed by the National Institute of Health as part of their evidence-based cancer control programs, that promote HPV vaccination education in various communities (16).

Our findings differed from a prior data analysis from the 2013-2014 National Health and Nutrition Examination Survey that showed HPV prevalence of any and high-risk genital HPV for adults aged 18–59 were 45.2% and 25.1% in men and 39.9% and 20.4% in women, respectively (17). Our results, conversely, showed a greater HPV infection prevalence among women than men. This could be explained by differences in the length of time studied or due to obtaining information from hospital discharges versus an examination survey (18, 19).

Differences in HPV diagnoses were also observed among various racial groups. Whites had the highest number of hospitalizations associated with an HPV diagnosis, followed by Hispanic individuals, then Blacks. Our estimate was similar to the study by Pierre-Victor, Trepka (20), which found a higher HPV prevalence among non-Hispanic White females in the United States between 14-59 years old. Some prior studies, however, revealed higher HPV infection rates among non-Hispanic Black adults in the United States. For instance, one study showed that Black women had twice the prevalence of HPV infection as other racial groups (21). The difference in our results could be that other racial groups are more likely to go to the hospital than Blacks. Due to our information being obtained from hospital discharges, this would have resulted in an underreported percentage of HPV infection among Blacks. The fact that the prevalence ratios of Blacks and Hispanics were higher than Whites in our results could be a further testament to this concept.

Disparities associated with HPV infection rates were seen in lower socioeconomic groups. Individuals with lower household incomes had a higher prevalence of HPV infections, while higher incomes were associated with the lowest number of infections diagnosed. An association was also found in the primary insurance of HPV-infected individuals, with most of those diagnosed with HPV infection having Medicaid insurance as the primary payer. This correlates with other studies that found low socioeconomic status as an associated risk factor for HPV infection. When looking at a group of low-income, diverse young women, Shikary, Bernstein (22) found the prevalence of HPV infection to be 68%, much higher than the national average of 45.2%. These results illustrate that patients with lower socioeconomic status are at an increased risk of HPV infection and should, therefore have increased resources for HPV education, vaccination, and screening.

Geographically, more infections were observed in the southern regions of the United States. Currently, the South has one of the lowest HPV vaccination rates in the USA (23). Moreover, the high-risk type of HPV infection is higher in the South than in other regions. (20). Benard, Thomas (24) found the highest levels of HPV-associated cervical cancer in Arkansas, Oklahoma, Alabama, Tennessee, and Texas. These results indicate that additional efforts are needed to address HPV prevention and early detection in the southern United States.

Vaccination is currently the most effective method to address the increasing incidence of HPV infection in the United States, and early screening is needed to detect HPV-related neoplastic transformation at its early stages (6). Additional research is needed to identify barriers to HPV vaccination and to investigate why HPV vaccination rates remain lower than other routine vaccinations. In addition, clinical trials are needed to determine how to achieve efficacious early screening for non-cervical, HPV-related cancers. Future studies should also investigate the most effective methods of disseminating information about HPV infection, vaccination, and screening to those with significant risk factors who are unlikely to attend regular wellness visits for early screening and vaccination.

### Limitations

A few significant elements in this study’s design and analysis could result in misunderstanding. No longitudinal data are available due to NIS’s inability to identify specific patients; ergo, numerous recordings may be made because of the same patient’s repeated hospitalizations. Thus, it is important to understand that the epidemiologic data presented shows the trends and characteristics leading to hospitalization associated with a primary or secondary diagnosis of HPV, not the trends and characteristics of becoming infected with HPV. Furthermore, NIS data does not consider outpatient interactions and observation-only stays, so there may be an underrepresentation of patients infected with HPV who are being monitored only in an outpatient setting. In addition, the sampling methodology forbids the estimation of volumes for subsets (for example, state-level estimates cannot be derived by applying discharge weights to all sampled hospitals/patients within a state). Finally, the administrative, diagnostic, and procedural codes utilized by NIS for the linked hospitalizations are supplied on the billing record. They are not individually validated due to changes in methodology in 2012 that prevented hospital-level estimates from being produced.

### Conclusion

Overall, the prevalence of HPV has increased in the United States in the past 12 years. Understanding the trends and characteristics of HPV infection can contribute to public health interventions aimed at prevention and early detection. Efforts to improve HPV vaccination rates and address risk factors associated with HPV infection are crucial to reducing the burden of HPV-related diseases. Further research can be conducted among those at higher risk to investigate the underlying factors responsible for the disparities and increasing rates of HPV observed in this study.

## Appendices

**Appendix A: ICD-9 and ICD-10 code description for HPV:**

ICD-9 codes

795.05 = cervical high risk human papillomavirus; DNA test positive

795.09 = other abnormal Papanicolaou smear of cervix and cervical human papillomavirus; cervical low-risk human papillomavirus; DNA test positive.

796.75 = anal high risk human papillomavirus; DNA test positive

796.79 = other abnormal Papanicolaou smear of anus and anal HPV; anal low-risk human papillomavirus; DNA test positive.

ICD-10 codes

R87.810 = high-risk human papillomavirus DNA test positive R87.820 = cervical low-risk human papillomavirus DNA test positive

R85.81= high-risk human papillomavirus DNA test positive from female genital organs R85.82 = low-risk human papillomavirus DNA test positive from female genital organs

## Data Availability

All relevant data are within the manuscript and its supporting information files.

